# Pulmonary function and risk of Alzheimer dementia: two-sample Mendelian randomization study

**DOI:** 10.1101/2020.07.28.20163352

**Authors:** Tom C. Russ, Sarah E. Harris, G. David Batty

## Abstract

Dementia is a major global public health concern and in addition to recognised risk factors there is emerging evidence that poorer pulmonary function is linked with subsequent dementia risk. However, it is unclear if this observed association is causal or whether it might result from confounding. Therefore, we present the first two-sample Mendelian randomisation study of the association between pulmonary function and Alzheimer dementia using the most recent genome-wide association studies to produce instrumental variables for both. We found no evidence of a causal effect of reduced Forced Expiratory Volume in 1 second (FEV_1_) or Forced Vital Capacity (FVC) on Alzheimer dementia risk (both P>0.35). However, the FEV_1_/FVC ratio was associated with Alzheimer dementia risk with, in fact, superior function predicting an increased dementia risk (OR 1.12, 95%CI 1.02-1.23; P=0.016) which may result from survivor bias. While we can conclude that there is no causal link between impaired pulmonary function and Alzheimer dementia, our study sheds less light on potential links with other types of dementia.

---

Dementia is a major growing global public health problem.^1^ Alzheimer disease risk is thought to be raised in the presence of relatively few environmental and genetic factors including lower educational attainment, hypertension, obesity, diabetes, cigarette smoking, and the *APOE* ε4 allele.^2^

Recent findings also suggest that impaired pulmonary function is consistently associated with ∼40% elevation in later dementia risk.^3^ While there is mechanistic evidence to support this — including hypoxia from extended sub-optimal ventilatory function^4^ — crucially, given the observational nature of these studies, it is unclear if this relationship is causal. An obstacle to drawing causal inference from such studies is the perennial problem of confounding — that characteristics of people poorer pulmonary function differ from the unexposed in various ways that may explain the association. Investigators attempt to include as many relevant covariates as possible but the possibility of confounding by unmeasured/imprecisely quantified factors is universal. Mendelian randomisation (MR) has been seen as a possible remedy to this problem^5^ and has been extended to two-sample MR where genetic associations for the exposure and outcome are obtained from independent samples.^6^ Accordingly, for the first time to our knowledge, we present a two-sample MR study to clarify whether the observed association between poorer pulmonary function and subsequent Alzheimer dementia (AD) is causal.

## METHODS

We ran a two-sample MR using summary data from the UKBiobank/SpiroMeta Consortium Genome-Wide Association Study (GWAS) comprising 400,102 individuals.^7^ We derived two genetic instruments for lung function: Forced Expiratory Volume in one second (litres; FEV_1_) and Forced Vital Capacity (litres; FVC). Of the 279 SNPs associated with lung function, but not smoking, only those related to the relevant trait with P<5×10^−8^ and the same direction of effect in UKBiobank and SpiroMeta were used as genetic instruments. In addition, we included a more exploratory measure: the FEV_1_/FVC ratio — which has been used in the diagnosis of chronic obstructive pulmonary disease whereby lower values are more suggestive of this condition.^8^ For the outcome we used summary data from the most recent GWAS which included 21,982 people with AD and 41,944 controls.^9^ The models used the TwoSampleMR R package.^10^ Since this study used publicly available data, no ethical approval was required.

## RESULTS

**Table 1** shows the relationship between lung function and subsequent AD risk. There was no evidence of a causal effect of poorer lung function — using FEV_1_ or FVC — on AD risk (both P>0.35). However, each SD increase in FEV_1_/FVC ratio (indicating superior lung function) was associated with an increased AD risk (OR,95%CI 1.12,1.02-1.23;P=0.016). The MR Egger intercept for the latter indicates little horizontal pleiotropy (**β**=0.0002,P=0.96) and the inverse-variance weighted Q-value (177.7,P=0.08) suggests no substantial heterogeneity. Using the weighted median method gave a similar result (1.15,1.00-1.31;P=0.048).

## DISCUSSION

We found that the observed association between lower pulmonary function and AD risk was not supported as being causal. Thus, it is possible that the original relationship resulted from confounding by one or more unmeasured/poorly measured confounders. Multiple candidates exist including an adverse intrauterine environment leading to reduced maximal lung function, exposure to environmental factors (e.g., tobacco smoke, atmospheric pollution) affecting lung function and development, and socioeconomic factors (poverty, educational failure, and less-advantaged social class). In our systematic review and meta-analysis, most included studies took account of smoking and cardiovascular disease risk factors, and slightly fewer included height.^3^ Socioeconomic position was variably accounted for and there was little coverage of the whole life course in terms of all included covariables.

**Table 1.**
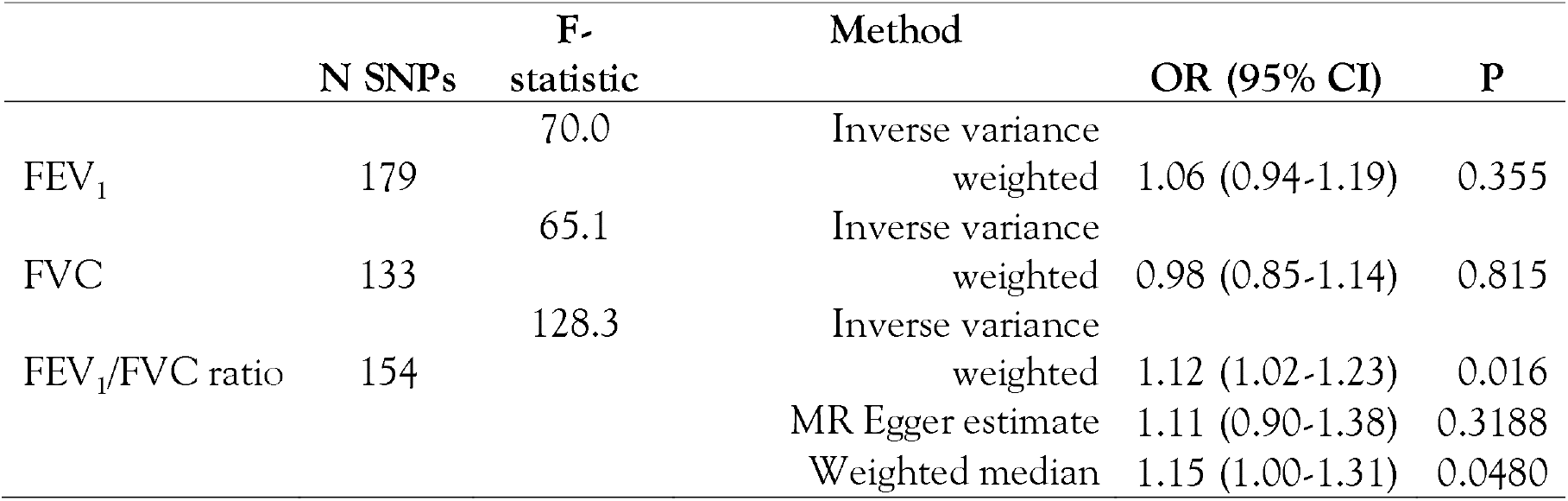
Estimates of the association between pulmonary function (FEV_1_, FVC, and FEV_1_/FVC ratio) and Alzheimer dementia from a two-sample Mendelian randomization

The FEV_1_/FVC ratio has not been routinely examined in relation to dementia risk.^3^ However, we found a link — albeit a weak one — between higher pulmonary function captured by this measure and increased AD risk. This may possibly be explained by survivor bias, with participants with poorer pulmonary function dying before they reach late life, but a false positive result must also be considered.

MR uses genetic variants which are randomly allocated at conception — and therefore generally independent of confounders that may otherwise bias an association when using observational methods — as proxies for environmental exposures. This assumes that genetic variants are:

(1) associated with the exposure; (2) only associated with the outcome of interest via their effect on the exposure; and 3) independent of confounders. It also relies on the exposure being accurately measured in the GWAS from which the instrument is derived. Pulmonary function was accurately measured with rigorous quality control in both UKBiobank (87.2% participants) and the individual studies of the SpiroMeta consortium.^7^ Pathway analysis suggested biological plausibility for the SNPs used as instruments with enrichment of genes relating to extracellular matrix organisation and ciliogenesis.^7^ Furthermore, a genetic risk score comprising all 279 lung function SNPs predicted COPD.^7^ It is unlikely that collider bias due to smoking and height adjustment in the lung function GWAS explains the observed association, as SNPs associated with smoking behaviour were excluded and a sensitivity analysis excluding the 12 SNPs included in our instrument which were associated with height in UKBiobank did not affect our conclusions.

The AD GWAS included 46 case-control studies from four consortia; rates of *APOE* e4 carriage are not reported.^9^ These studies used various methods of ascertaining dementia, with multiple diagnostic criteria being applied. Some studies used clinical diagnoses and some identified Alzheimer-type pathology post mortem. This variation is likely to affect the applicability of the GWAS findings in our analysis.

In contrast to the instrumental AD variable used here, most observational studies use a more general category of ‘dementia.’^3,9^ This lack of clarity is common and the multiple diseases causing the dementia syndrome — e.g., Alzheimer disease, cerebrovascular disease, Lewy body disease, and Fronto-Temporal Lobar Degenerative syndromes — are frequently conflated. Depending on the methodology used, clarifying an individual’s precise diagnosis can be challenging. For example, death certificates frequently only record the broad dementia syndrome. Thus, while we can conclude that there is no causal link between impaired pulmonary function and AD, our study sheds less light on potential links with other types of dementia. It is plausible that there may be a different relationship between pulmonary function and vascular dementia, for instance.

## Data Availability

All data are publicly available.

